## Supplementary Information for "A blood- and brain-based EWAS of smoking"

**Blood- and Brain-Based Epigenome-Wide Association Studies of Smoking**

**Supplementary Information**

**Supplementary Figures**

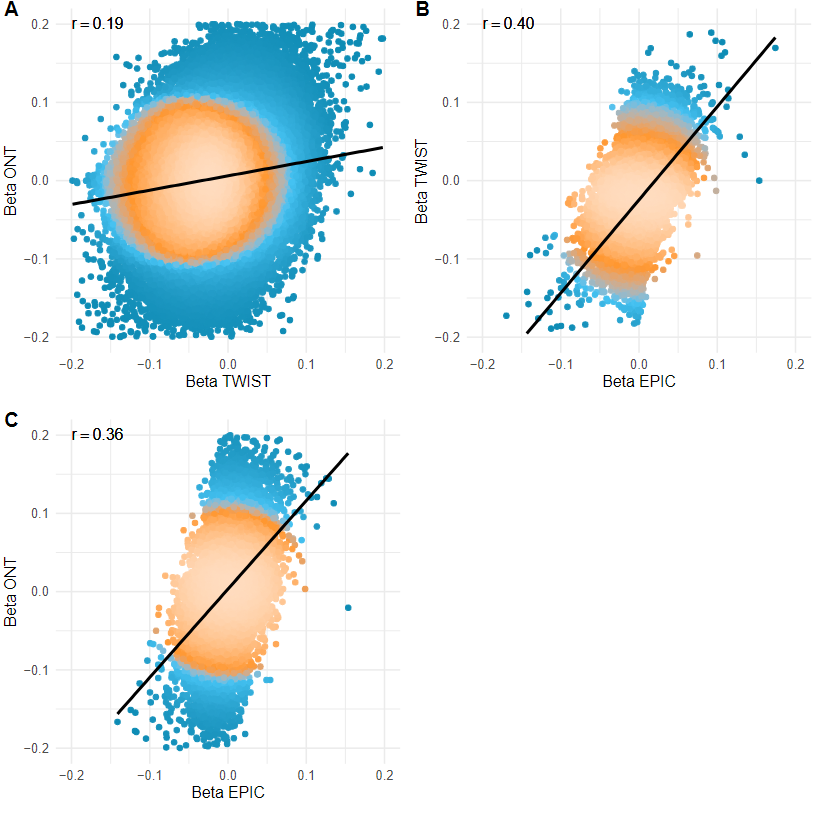

**Supplementary Fig. 1. Comparison of beta estimates from various Epigenome-Wide Association Studies (EWASs) of smoking (n=46 individuals). Methylation data were collected using three platforms: the EPIC array, the TWIST Human Methylation Kit (short-read targeted DNA sequencing), and Oxford Nanopore Technologies (ONT, long-read DNA sequencing).** The black line represents a fitted linear regression, while points are color-coded by density, ranging from blue (least dense) to bright yellow (most dense). The value r corresponds to Pearson’s correlation coefficient. Data were filtered to include only points significant at P < 0.05 in at least one dataset before plotting.

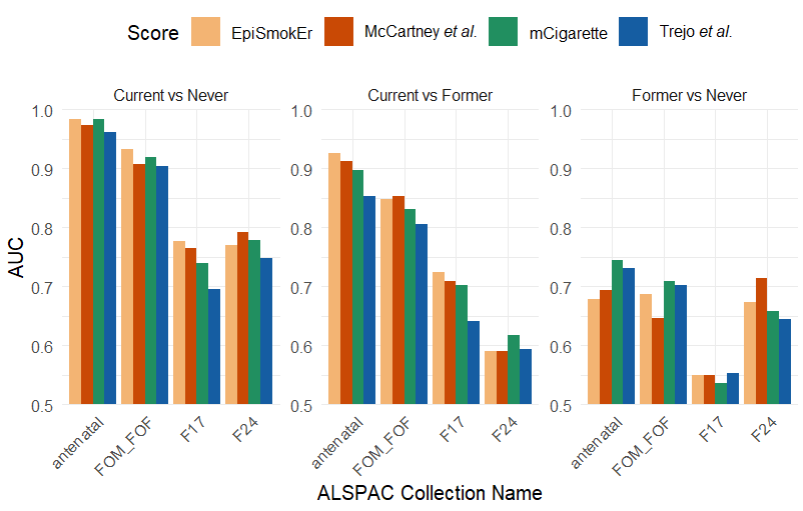

**Supplementary Fig. 2. The performance (measured by Area Under the Curve (AUC)) of four smoking-related epigenetic scores (EpiSmokEr, McCartney et al.** ^1^**, mCigarette, and Trejo et al.** ^2^**) across different ALSPAC data collection time points and smoking status comparisons.** Source data and the number of samples used to derive statistics are provided as a Source Data file.

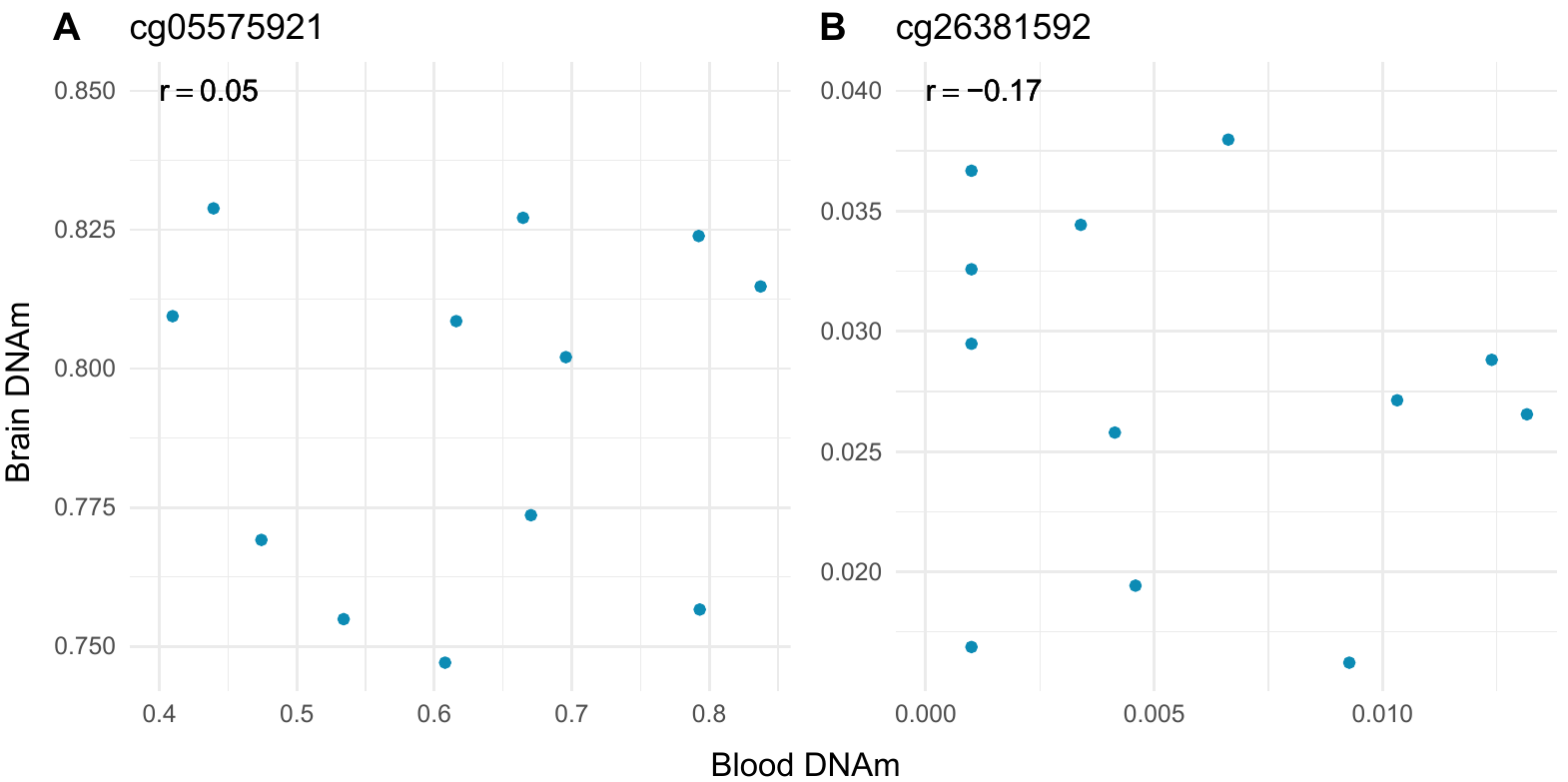

**Supplementary Fig. 3. DNA methylation levels were measured at two CpG sites (A: cg05575921, B: cg26381592) in blood (latest available sample) and hippocampus tissue.** For one participant of Lothian Birth Cohort 1936, only a hippocampus sample was available (n=13 samples), with no corresponding blood sample (n=12 samples). The value of r indicates the Pearson’s correlation coefficient. Source data are provided as a Source Data file.

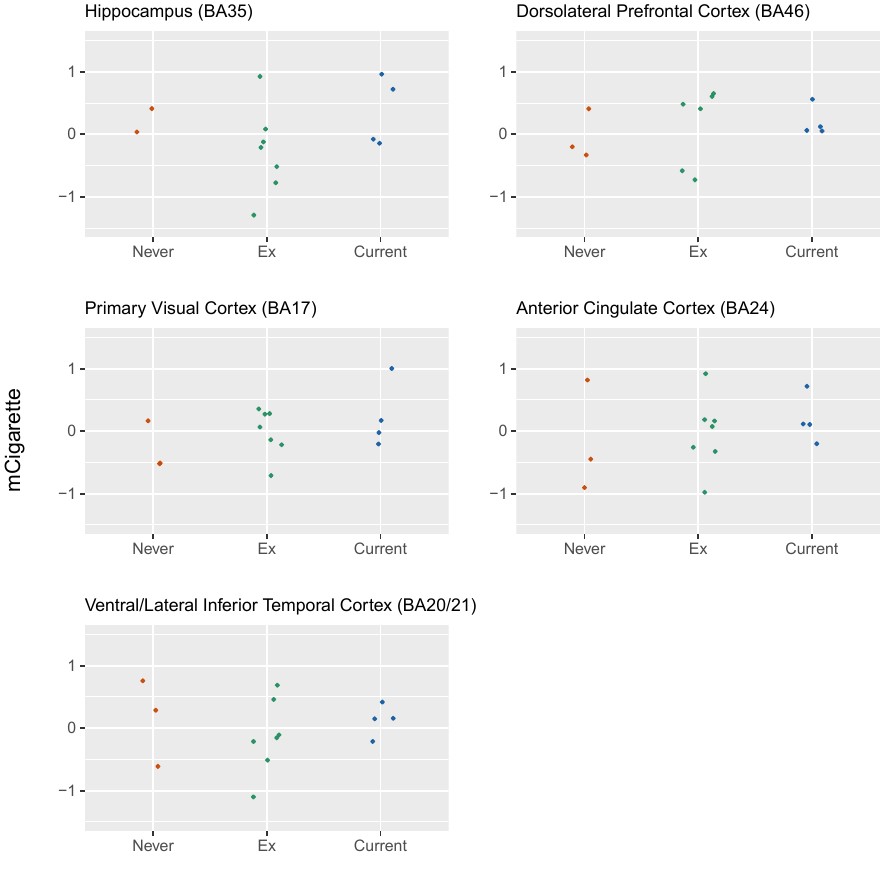

**Supplementary Fig. 4. mCigarette applied to post-mortem brain samples from** **Lothian Birth Cohort 1936 (n=14 samples, n_hippocampus_=13 samples).** Three smoking categories were considered (never, former and current smokers). The association was plotted across five brain regions. Source data are provided as a Source Data file.

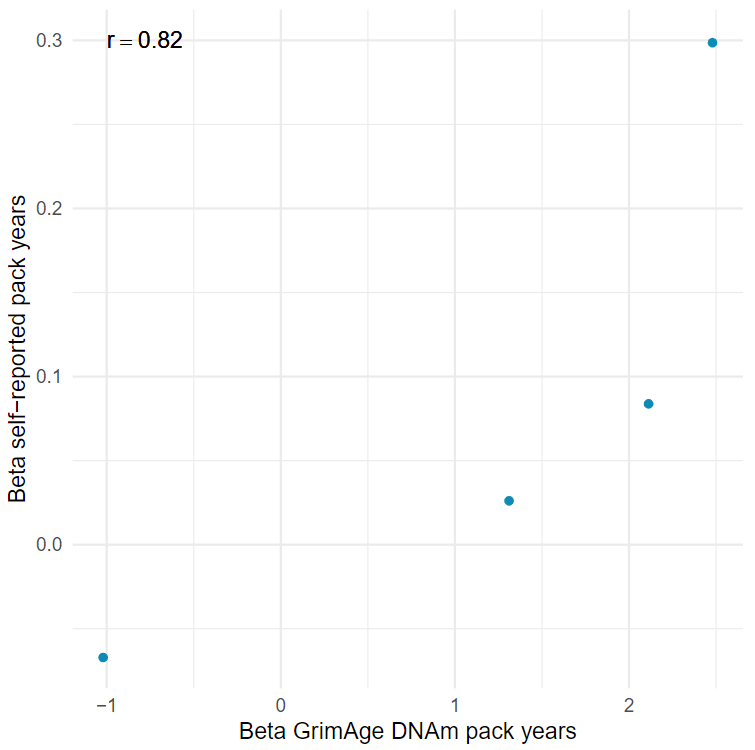

**Supplementary Fig. 5. Comparison of beta coefficients for lead loci identified in the GrimAge DNAm pack years Genome-Wide Association Study (GWAS) and the smoking pack years GWAS.** Both studies were conducted in Generation Scotland (n=17,105 individuals). Blue dots represent the lead loci. The r corresponds to the Pearson correlation coefficient. Source data are provided as a Source Data file.

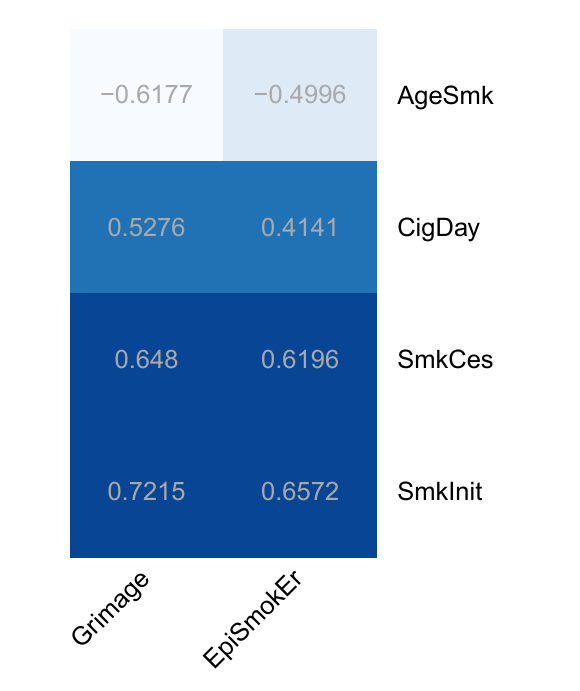

**Supplementary Fig. 6. Genetic correlations between epigenetic (EpiSmokEr, Grimage pack years) and self-reported smoking.** Genome-Wide Association Studies of EpiSmokEr and Grimage pack years scores were conducted in Generation Scotland (n=17,105 individuals). The results were compared to previously published GWAS of tobacco use (n_max_=2,669,029 Europeans), which considered four smoking behaviours: smoking initiation (SmkInit), smoking cessation (SmkCes), age of initiation of smoking (AgeSmk) and cigarettes per day (CigDay) (Saunders *et al.*, 2022) ^3^.

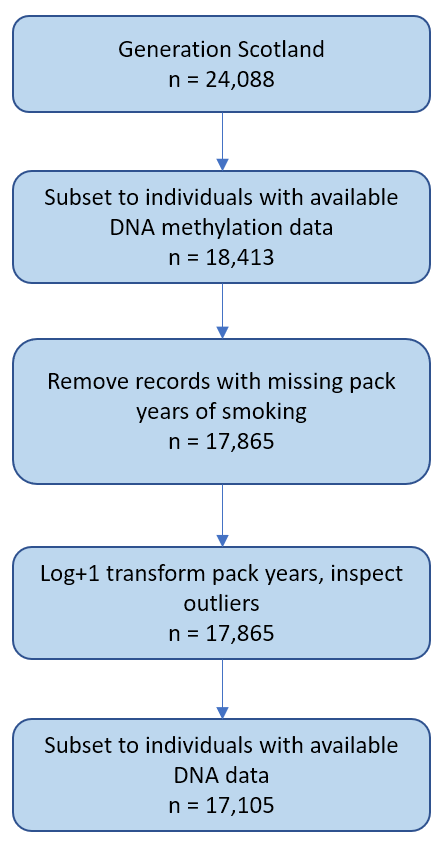

**Supplementary Fig. 7. Pack years data pre-processing in Generations Scotland.** Trimming outliers consisted of removing log+1 transformed pack years values more than 4 SDs from the mean. The sample size (n) represents the number of individuals.

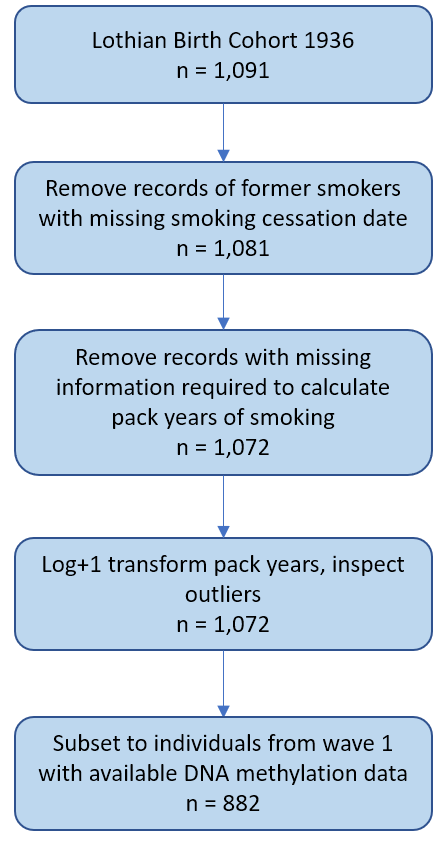

**Supplementary Fig. 8. Pack years data pre-processing in Lothian Birth Cohort 1936.** Records of participants that described themselves as never- or ex-smokers, provided the date they started smoking and did not provide the date they quit smoking were likely to contain data entry errors and were therefore removed from the dataset. Trimming outliers consisted of removing log+1 transformed pack years values more than 4 SDs from the mean. The sample size (n) represents the number of individuals.

**Supplementary Methods**

**Sample Selection for Generation Scotland – TWIST**

**Objective:** Select 24 current smokers (cases) and 24 age- and sex-matched never smokers (controls) from wave 3 in Generation Scotland.

**Protocol:** We focused on individuals from wave 3 of Generation Scotland given that this wave contains unrelated individuals (n=4,450). We then stratified wave 3 individuals by self-reported smoking behaviour at baseline as follows: 675 current smokers (possible cases), 1,398 ex-smokers (excluded), 101 with missing data (excluded) and 2,276 never smokers (possible controls).

*Case definition:* We then examined the distribution of smoking pack years among the 675 current smokers (or possible cases). Our goal was to identify individuals with high smoking consumption in order to ensure a robust methylation signal would be present when comparing cases against controls. Figure 1 shows the distribution of smoking pack years among these individuals. Smoking pack years were transformed by using a log(units+1) transformation to reduce skew. We selected individuals in the range 4.0-4.5 on the log(pack years+1) scale, corresponding to a range of 53.9-87.7 pack years. There were 40 current smokers (possible cases) in this range.

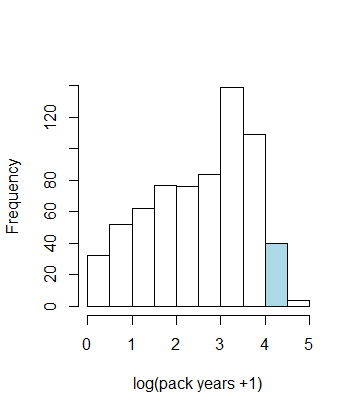

**Figure 1. Selection of current smokers (n=675 possible cases) based on high smoking consumption in wave 3 of Generation Scotland.** Forty selected individuals are highlighted in blue.

*Case and control selection:* In total, 12 males and 12 females were randomly sampled from the pool of 40 possible cases. The R package *Matchit* was then used to select 24 age- and sex-matched controls from the pool of 2,276 never smokers.

*Diagnostics:* We then examined whether cases and controls showed a clear separation in terms of their methylation at the *AHRR* CpG probe cg05575921. Hypomethylation of this CpG site is strongly associated with smoking. Therefore, we expected that cases would have lower levels of methylation at this site. This was confirmed upon visual inspection of the diagnostic plot shown in Figure 2.

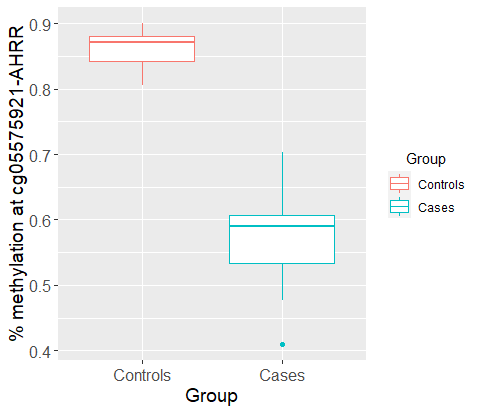

**Figure 2. Hypomethylation of AHRR probe in smokers versus non-smokers.** There was clear separation of methylation levels at the smoking-associated probe cg05575921. Cases (current smokers, n=24 individuals) are shown in blue and controls (never smokers, n=24 individuals) are shown in red. Box plots are defined as follows: the centre line represents the median (50th percentile). The box bounds indicate the interquartile range (IQR; 25th to 75th percentile). Whiskers extend to the smallest and largest values within 1.5×IQR.

Lastly, we performed linear epigenome-wide association studies on (i) smoking pack years (continuous trait) and (ii) case-control status (binary trait) using OSCA software. Models were adjusted for age and sex. The smoking-associated probe cg05575921 was the strongest correlate of both traits with *P*<10^-20^. The next most significant CpGs were also known smoking correlates, confirming that we are able to see clear biological signals within the sample of 24 cases and 24 controls selected in our protocol.

**Summary of Sequencing Protocol - Oxford Nanopore Sequencing**

Forty-eight Generation Scotland DNA samples were selected and assessed for quality using the Fragment Analyzer (Agilent) with the Genomic DNA 50kb Kit to evaluate size and integrity, and quantified by Qubit 2.0. Purity was checked via Nanodrop-8000. Each DNA sample (1.5 µg) underwent purification, repair, end-prepping, and barcoding with the ONT Native Barcoding Kit 24, alongside NEBNext modules for ligation and repair. Libraries were prepared without DNA shearing for the initial 24 samples; subsequent samples were sheared to a target of 10kb. All samples were purified with AMPure XP beads, quantified, and pooled based on sex and age. After adapter ligation, sequencing libraries were enriched for fragments longer than 3kb. Libraries were quantified with the Qubit dsDNA HS assay. Sequencing was conducted on the Oxford Nanopore PromethION 24 with R10.4.1 flow cells, running for 72 hours. The first 24 libraries were sequenced at Edinburgh Genomics without basecalling, with later libraries basecalled on the Genetics Core PromethION. The following run settings were applied: 72h limit, active channel selection, pore scan every 1.5h, minimum read length of 200bp. MinKNOW software managed data acquisition, real-time feedback, basecalling, and device control. Dorado, optimized for NVIDIA GPUs, was used for high-accuracy basecalling, alignment, and modified base detection. Basecalling and demultiplexing were conducted on the Genetics Core PromethION server with modified bases set to detect CpG contexts (5mC and 5hmC).

**Supplementary References**

1.	McCartney, D. L. *et al.* Epigenetic prediction of complex traits and death. *Genome Biology* **19**, 136 (2018).

2.	Trejo Banos, D. *et al.* Bayesian reassessment of the epigenetic architecture of complex traits. *Nat Commun* **11**, 2865 (2020).

3.	Saunders, G. R. B. *et al.* Genetic diversity fuels gene discovery for tobacco and alcohol use. *Nature* **612**, 720–724 (2022).
